# Preferences for PrEP service delivery among adolescent girls and young women in remote villages in Lesotho: a discrete choice experiment

**DOI:** 10.64898/2026.05.27.26352981

**Authors:** Andréa Williams, Michael Strauss, Ottavia Prunas, Felix Gerber, Fabian Raeber, Giuliana Sanchez-Samaniego, Elis Saavedra, Tamaryn Crankshaw, Gavin George, Moruthoane Motlalentoa, Limakatso Mofilikoane, Mosa Mohasoa, Ravi Gupta, Mamoronts’ane Sematle, Makhebe Khomolishoele, Pauline Grimm, Irene Ayakaka, Tarumbiswa Tapiwa, Nthuseng Bridgett Marake, Rosina Phate-Lesihla, Maja Weisser, Alain Amstutz, Niklaus D. Labhardt

## Abstract

**Introduction:** Adolescent girls and young women (AGYW) in southern Africa are disproportionately affected by HIV. Despite increasing availability of HIV pre-exposure prophylaxis (PrEP), uptake and sustained use remain low. Existing service delivery models may not adequately meet the needs of AGYW, particularly in remote settings. We conducted a discrete choice experiment (DCE) to assess preferences for PrEP service delivery among AGYW living in Lesotho, a country with one of the highest HIV incidence rates globally.

**Methods:** The DCE was conducted among AGYW (16-24 years) in two districts in Lesotho. Participants completed a series of binary choice tasks comparing hypothetical PrEP service delivery scenarios defined by six attributes: service location, provider type, provider characteristics, provider confidentiality, PrEP product type, and the combination of additional prevention services offered. Preferences were analysed using mixed logit and latent class models.

**Results:** A total of 537 AGYW (median age 19 years, IQR 17–22) were included. Provider confidentiality was the strongest driver of choice, with non-confidential providers significantly less preferred (β = −0.58; 95% CI −0.69 to −0.46). Compared with nurses, services delivered by trained CHWs were preferred (0.17; 0.01 to 0.33), while those provided by doctors were less preferred (−0.15; −0.30 to 0.00). Younger female providers were preferred over older female providers (0.20; 0.04 to 0.36). Compared with the daily oral pill, both the 2-monthly injectable (−0.24; −0.39 to −0.08) and the vaginal ring (−1.02; −1.20 to −0.82) were less preferred. Differences in preferences were observed across age groups and districts. Latent class analysis identified two preference profiles, indicating variation in preferences for delivery and product characteristics.

**Conclusions:** Preferences for PrEP delivery among AGYW in Lesotho were strongly influenced by provider confidentiality. Among some AGYW, there was openness to decentralised delivery, particularly through CHWs and community-based models, which may reduce access barriers in remote settings. Product preferences were varied, and not all options were acceptable. Differences by age group and district indicate that no single delivery model will meet all needs. Building on the current standard of care, offering acceptable options in accessible and confidential ways may support PrEP uptake.

## Introduction

Across southern and eastern Africa, adolescent girls and young women (AGYW) are disproportionately affected by HIV [1]. Lesotho has one of the highest HIV incidence and prevalence rates globally (0.64% per year and 27.4%), with prevalence reaching 13% among those aged 20–24 years [2]. As in other countries in the region, AGYW are a priority population for HIV prevention [3,4].

HIV pre-exposure prophylaxis (PrEP) is a highly effective prevention option for individuals at increased risk of HIV; however, uptake and sustained use remain low in many high-prevalence settings [5,6]. Low PrEP use among AGYW is a recognised challenge across southern Africa [7,8]. In Lesotho, PrEP is primarily delivered as daily oral tenofovir-based medication through nurse-led public-sector health facilities; however, existing delivery models may not adequately engage AGYW [9,10]. Decentralised and community-based delivery models, as well as long-acting formulations such as lenacapavir, present promising opportunities, but remain limited in availability and are not yet implemented at scale.

In remote areas of Lesotho, structural barriers limit access to PrEP, including long travel distances, inconvenient clinic hours, and limited availability of services in understaffed facilities [10]. Community-based approaches, such as mobile clinics and community health workers (CHWs), offer strategies to extend delivery beyond facility settings. Evidence from similar contexts suggests that these models can improve access to HIV prevention and sexual and reproductive health services, including PrEP [11–13].

However, improving access alone may be insufficient to increase PrEP uptake and sustained use among AGYW. Decisions about PrEP use are influenced by a range of service delivery and product-related factors [14–17]. Designing interventions that reflect these preferences is important for improving initiation and sustained use. To inform PrEP service design and delivery, we conducted a discrete choice experiment to assess preferences for PrEP products and service delivery among AGYW in Lesotho.

## Methods

### Study setting

We conducted a DCE within the Community-Based Chronic Care Lesotho (ComBaCaL) population-based cohort in the northeastern districts of Butha-Buthe and Mokhotlong. Data were collected between September and December 2024. These districts include remote mountain villages with limited access to health facilities. Within each village, Ministry of Health-trained CHWs provide basic health services and facilitate linkage to facility-based care, with a subset receiving additional training and support through the ComBaCaL program [19].

### DCE concept and design

A DCE is a stated-preference method used to understand how individuals make choices when presented with hypothetical but realistic scenarios [20]. DCEs are particularly useful where real-world choices are limited or not directly observable, and where it is important to assess preferences across competing service features.

In this study, participants completed a series of binary choice tasks comparing two hypothetical PrEP service delivery scenarios defined by different combinations of attributes. Participants selected their preferred option in each task. A forced-choice, unlabeled design was used, in which alternatives were presented as combinations of attributes (‘Option A’ and ‘Option B’) rather than predefined service models. No opt-out option was included, to focus on relative preferences between attributes.

Six attributes were included: service location, provider type, provider characteristics (age and sex), provider confidentiality, PrEP product type, and the combination of additional prevention services offered (i.e. contraception and STI screening). Each attribute included multiple levels representing different ways the service could be delivered, and combinations of these levels were presented across the choice sets. The study was designed and reported in accordance with the DIRECT checklist for discrete choice experiments [21]. An example of a choice set is shown in Figure 1.

**Figure 1.**
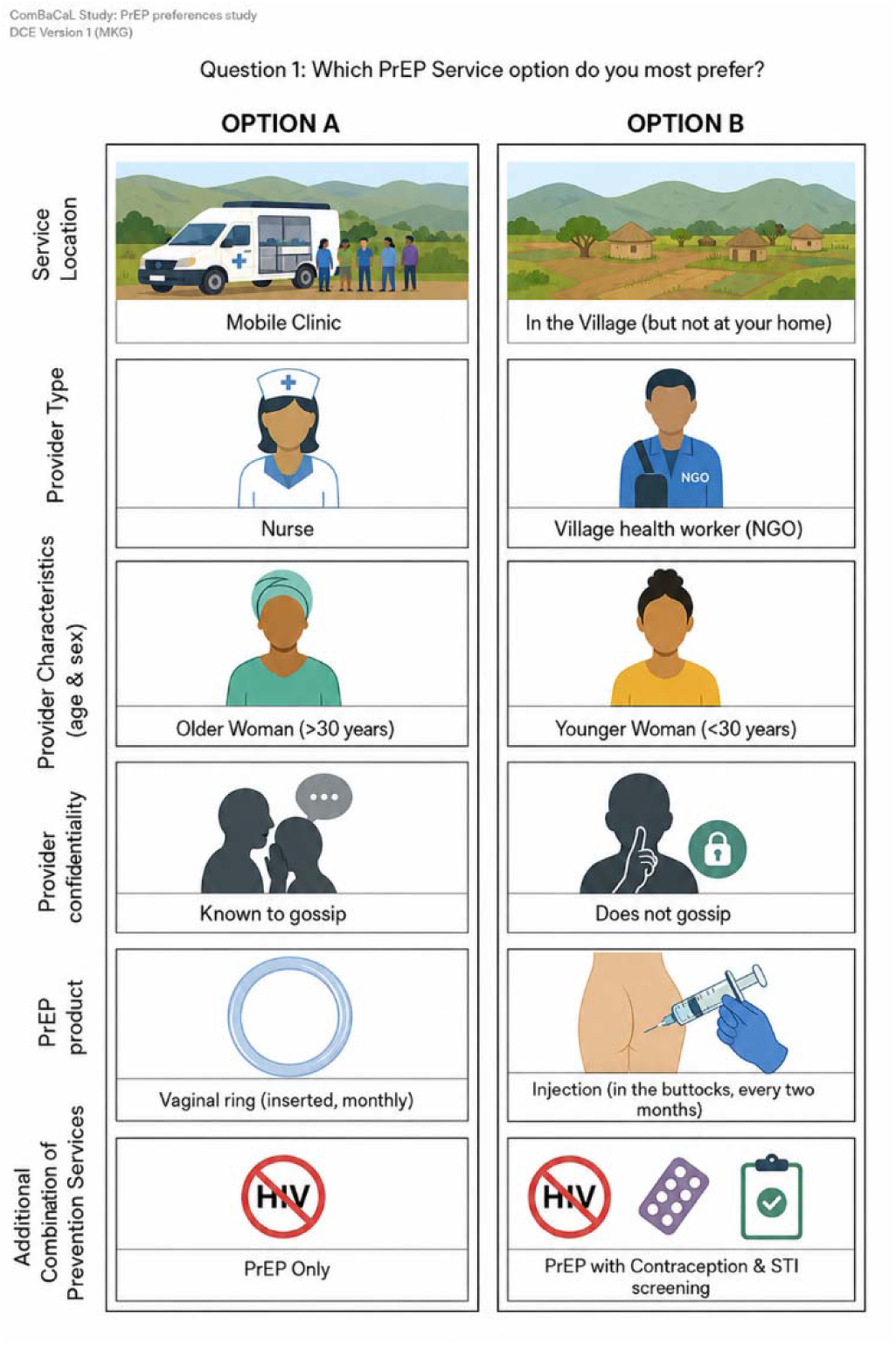
Example of a discrete choice experiment (DCE) set comparing two hypothetical pre-exposure prophylaxis (PrEP) service delivery options.□

### Attribute development and selection

Attributes and levels were identified through a combination of literature review, formative qualitative research, and consultation with HIV prevention experts and local stakeholders. Service location, provider type, PrEP product type, and additional services were informed by previous discrete choice experiments and HIV prevention literature among AGYW in southern Africa [16,17]. Additional attributes were informed by formative qualitative research conducted among AGYW in the study districts. Focus group discussions highlighted provider confidentiality and provider characteristics, particularly age (with a cut-off of 30 years to distinguish between older and younger providers) and sex, as important factors influencing trust and comfort when accessing HIV prevention services.

The final attributes, levels, and reference categories are presented in Table 1, with the full set provided in Supplementary Tables S1a and S1b.

**Table 1.** Final attributes and levels included in the discrete choice experiment design.

| Attribute | Levels |
| --- | --- |
| Service location | Home (in the village); **Healthcare facility**□; Mobile clinic; In the village (but not at home) |
| Provider type | Doctor; **Nurse**□; CHW (trained); CHW (routine) |
| Provider characteristics | Younger male (<30 years); Younger female (<30 years); Older male (>30 years); **Older female (>30 years)**□ |
| Provider confidentiality | Non-confidential; **Confidential**□ |
| PrEP product | **Oral pill (daily)**□; Injection in buttocks (every 2 months); Injection in abdomen (every 6 months); Vaginal ring (monthly) |
| Combination of additional prevention services | **PrEP only**□; PrEP & contraception; PrEP & STI screening; PrEP, contraception & STI screening |

□ Levels used as the reference category for the analysis, selected to correspond to the current standard of care.

Some attributes commonly included in DCEs were excluded. Cost was not included, as PrEP is provided free of charge in Lesotho [3]. Side effects were excluded as they relate to specific PrEP formulations rather than service delivery models. Instead, formulation-related characteristics likely to influence preferences and delivery feasibility, including method of administration and dosing frequency, were captured under the ‘product type’ attribute. Waiting time and travel distance were excluded as they could not be applied consistently across household- and village-based delivery scenarios, and access burden was already captured by the ‘service location’ attribute.

### Experimental design and blocking

A fractional factorial, D-efficient design was generated in Stata (version 18) [22–24] to maximise statistical efficiency while minimising participant burden (Supplementary File S1). Choice sets were divided into six versions, with participants sequentially allocated to one version during recruitment. Each participant completed 10 choice tasks, including a repeated (dominant) choice set to assess response consistency (Supplementary File S2).

### Sample size estimation

Sample size estimation followed a commonly used convention for DCEs (22): N ≥ 500 × l / (J × S), where l is the largest number of attribute levels [4], J is the number of alternatives per choice set [2], and S is the number of choice sets per respondent [8] - a minimum sample size of 125 participants per subgroup.

Subgroup analyses included age (16-19 vs. 20-24 years) and district (Butha-Buthe vs. Mokhotlong). A recruitment target of 150 AGYW per subgroup was set to allow for 20% data loss, resulting in a total sample of 600 participants.

### Sampling, eligibility and recruitment

A total of 40 of the 103 ComBaCaL villages (19 in Butha-Buthe and 21 in Mokhotlong) were purposively selected based on the size of the AGYW population (minimum of 10 AGYW per village). AGYW aged 16–24 years were eligible if they self-reported being female and reported an HIV-negative or unknown HIV status. Written informed consent was obtained from participants aged ≥18 years, while those aged 16–17 years provided written assent alongside parental or guardian consent.

Eligible AGYW were identified through the ComBaCaL cohort database. CHWs conducted door-to-door recruitment during routine household visits, providing information about the DCE and inviting eligible participants to participate.

### Survey administration and data collection

The DCE was administered by trained CHWs. Self-reported HIV status and HIV risk were recorded, along with basic sociodemographic information and access to services. The DCE was conducted in Sesotho using a paper-based visual tool (Supplementary File S3). Participants indicated their preferred option verbally, and responses were recorded electronically using a tablet-based application. CHWs were blinded to attribute combinations within each choice set and were unable to edit responses after submission.

### Data quality and participant comprehension

Responses to a repeated choice set were used to assess internal consistency and participant comprehension and were excluded from the final analysis if inconsistent. Participant comprehension was also assessed by CHWs using a five-point Likert scale, and cross-tabulated with repeated-choice consistency (Supplementary Table S2).

### Statistical analysis

A mixed-effects logistic regression model with all attribute levels specified as random parameters was used to estimate preferences for PrEP service delivery. Models were estimated using simulated maximum likelihood with 500 Halton draws. Estimated coefficients (β) represent the direction and strength of preferences relative to the reference category, while standard deviations indicate preference heterogeneity across participants.

Attribute levels were binary coded (Supplementary File S4), with reference categories corresponding to the current standard of care for PrEP delivery in Lesotho. Additional models were estimated stratified by age group (16–19 vs 20–24 years) and district (Butha-Buthe vs Mokhotlong).

Latent class logit models were estimated to further explore preference heterogeneity. Models with two to four classes were assessed, with selection based on model fit (Akaike Information Criterion [AIC] and Bayesian Information Criterion [BIC]), mean posterior probability of class membership, and the interpretability of class-specific preference patterns.

All analyses were conducted in R (version 4.3.2) [25].

### Ethics approval

The ComBaCaL cohort study and substudies were approved by the National Health Research Ethics Committee of Lesotho (Ref. ID 210-2022 and 210-2022 Nested) and the Ethics Committee of Northwestern and Central Switzerland (EKNZ) (Ref. AO_2022-058, AO_2022-00074 and AO_2022-00077). Ethics approval for the DCE was additionally obtained from the University of KwaZulu-Natal Biomedical Research Ethics Committee (Ref. HSSREC/00006690/2024).

## Results

### Eligibility and enrolment

According to ComBaCaL cohort records, 689 AGYW were eligible for participation across the 40 villages, of whom 556 were approached. Fifteen declined participation or withdrew consent, and four were excluded due to inconsistent responses during the DCE, resulting in a final sample of 537 and an overall response rate of 81%.

Overall, 91% of participants were rated as having good or very good understanding of the choice tasks, indicating high comprehension across the sample (Supplementary Table S2).

### Participant characteristics

Among the 537 participants, 53% were aged 16–19 years. Participants were evenly distributed across the two districts (52% in Butha-Buthe and 48% in Mokhotlong). Key participant characteristics by district are presented in Table 2.

**Table 2.** Baseline characteristics of participants (n = 537)

| Characteristic | Overall<br>n = 537 | Butha-Buthe<br>n = 281 | Mokhotlong<br>n = 256 |
| --- | --- | --- | --- |
| **Age group** |  |  |  |
| 16-19 | 283 (53%) | 154 (55%) | 129 (50%) |
| 20-24 | 254 (47%) | 127 (45%) | 127 (50%) |
| **Healthcare access** |  |  |  |
| Difficult Access <sup>1</sup> | 316 (59%) | 231 (82%) | 85 (33%) |
| Easy Access | 221 (41%) | 50 (18%) | 171 (67%) |
| **Relationship status** |  |  |  |
| In a committed relationship <sup>2</sup> | 232 (43%) | 114 (41%) | 118 (46%) |
| Single | 305 (57%) | 167 (59%) | 138 (54%) |
| **Highest level of education completed** |  |  |  |
| No education | 13 (2%) | 3 (1%) | 10 (4%) |
| Primary school education | 136 (25%) | 45 (16%) | 91 (36%) |
| Secondary/high school education | 361 (67%) | 211 (75%) | 150 (59%) |
| Tertiary education | 27 (5%) | 22 (8%) | 5 (2%) |
| **HIV testing history** |  |  |  |
| Tested HIV negative more than 6 months ago | 117 (22%) | 76 (27%) | 41 (16%) |
| Tested HIV negative within the last 6 months | 330 (62%) | 159 (57%) | 171 (67%) |
| Unknown HIV status/Refused to say | 90 (17%) | 46 (16%) | 44 (17%) |
| **Self-perceived risk of acquiring HIV<sup>3</sup>** |  |  |  |
| High | 161 (30%) | 81 (29%) | 80 (31%) |
| Low | 367 (68%) | 198 (71%) | 169 (66%) |
| Unknown/Refused to say | 9 (2%) | 2 (1%) | 7 (3%) |

<sup>1</sup>defined as needing to cross a mountain or river or travel >10 km to the nearest health facility
<sup>2</sup>Includes participants who were seriously dating, cohabiting, or married
<sup>3</sup>defined as self-reported unprotected sex in the past six months with one or more partner(s) whose HIV status is unknown

### Preferences for PrEP service delivery: main effects model

Results from the mixed-effects logistic regression model are presented in Figure 2. Services delivered by trained CHWs were preferred (β = 0.17; 95% CI 0.01 to 0.33), while services provided by doctors were less preferred (−0.15; −0.30 to 0.00), compared with nurses. Participants also preferred younger female providers over older female providers (0.20; 0.04 to 0.36).

**Figure 2:**
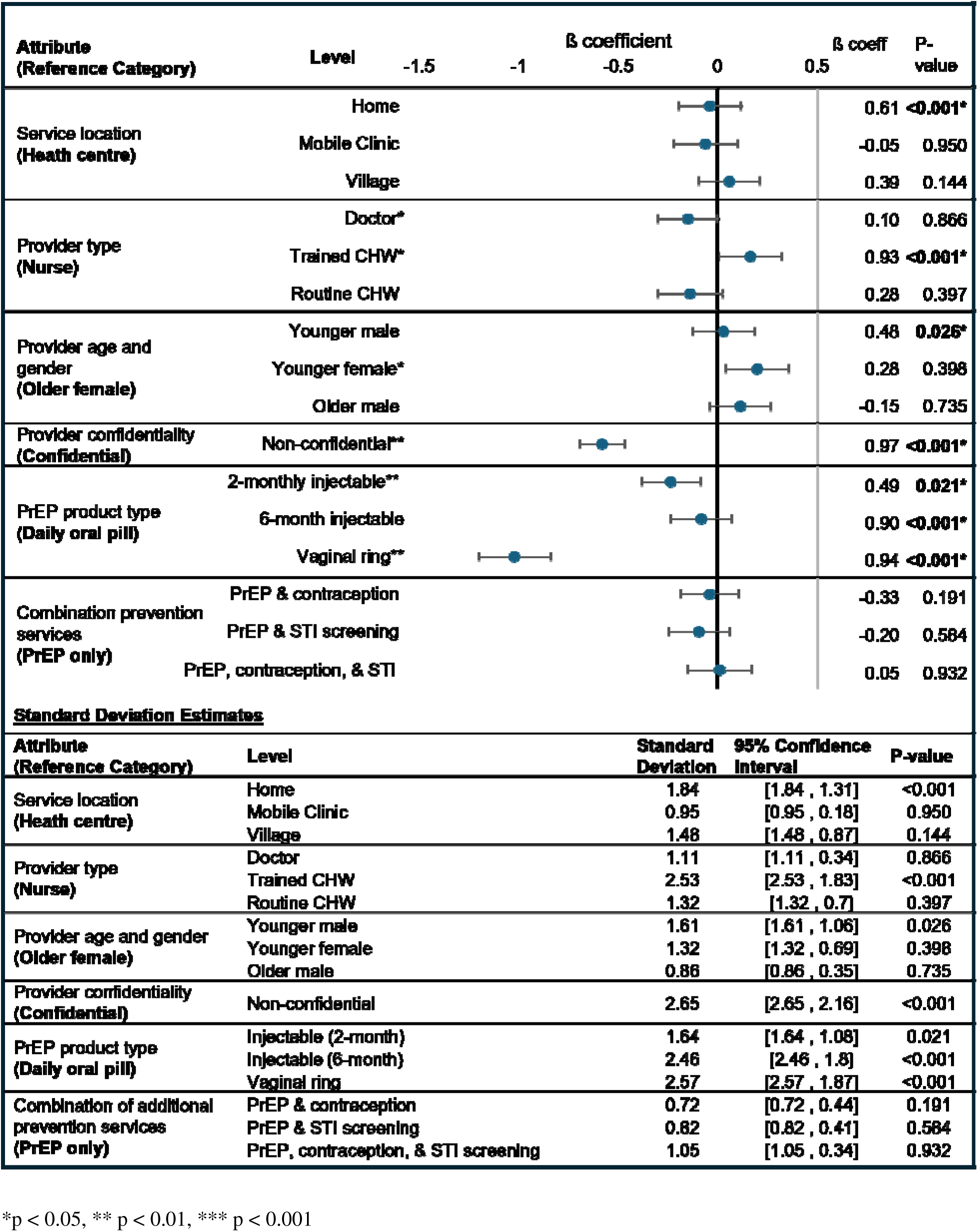
Main effects of PrEP service delivery preferences. beta coefficients and 95% confidence intervals

Provider confidentiality had a strong influence on choice, with non-confidential providers substantially less preferred (−0.58; −0.69 to −0.46). PrEP product type was also associated with preferences. Compared with the daily oral pill, the 2-monthly injectable PrEP (−0.24; −0.39 to −0.08) and vaginal ring (−1.02; −1.20 to −0.82) were less preferred, while no significant difference was observed for 6-monthly injectable PrEP (−0.08; −0.24 to 0.07).

Standard deviation estimates indicated significant variation in preferences across several attributes. Variation was observed for services delivered at home (SD 1.84; 95% CI 1.31–2.59) and by trained CHWs (SD 2.53; 95% CI 1.83–3.49), as well as for younger male providers (SD 1.61; 95% CI 1.06–2.45).

Preferences regarding provider confidentiality also varied considerably, particularly for non-confidential providers (SD 2.65; 95% CI 2.16–3.25). Significant variation was observed for PrEP product type, including the 2-monthly injectable (SD 1.64; 95% CI 1.08–2.51), 6-monthly injectable (SD 2.46; 95% CI 1.80–3.37), and vaginal ring (SD 2.57; 95% CI 1.87–3.53).

### Subgroup analyses of PrEP service delivery preferences by age group and district

Stratified mixed logit models by age group (16–19 vs 20–24 years) and district (Butha-Buthe vs Mokhotlong) showed patterns consistent with the overall model (Tables 3a-3b). Provider confidentiality and PrEP product type were strongly associated with choice across age groups and districts, with consistent aversion to non-confidential providers (−0.60; −0.78 to −0.43 in younger AGYW and −0.56; −0.73 to −0.39 in older AGYW) and the vaginal ring (−1.14; −1.47 to −0.84 and −0.97; −1.27 to −0.67, respectively). Differences emerged by age group. Mobile clinic services were less preferred among AGYW aged 16–19 years (−0.24; −0.46 to 0.00), with no difference observed among those aged 20–24 years. Routine CHWs were also less preferred among younger participants (−0.36; −0.58 to −0.13), while services delivered by trained CHWs were preferred among those aged 20–24 years (0.32; 0.08 to 0.56), with no clear preference among younger participants.

Differences were also observed by district. In Butha-Buthe, services provided by doctors were less preferred (−0.22; −0.45 to −0.01), while no difference was observed in Mokhotlong. Conversely, trained CHWs were preferred in Mokhotlong (0.34; 0.12 to 0.57), but not in Butha-Buthe. In Butha-Buthe, younger female providers (0.35; 0.11 to 0.59) and older male providers (0.32; 0.08 to 0.57) were preferred, while no differences were observed in Mokhotlong. In Mokhotlong, both the 2-monthly injectable PrEP (−0.32; −0.53 to −0.11) and 6-monthly injectable PrEP (−0.24; −0.45 to −0.02) were less preferred, whereas no differences were observed in Butha-Buthe.

**Table 3a.**
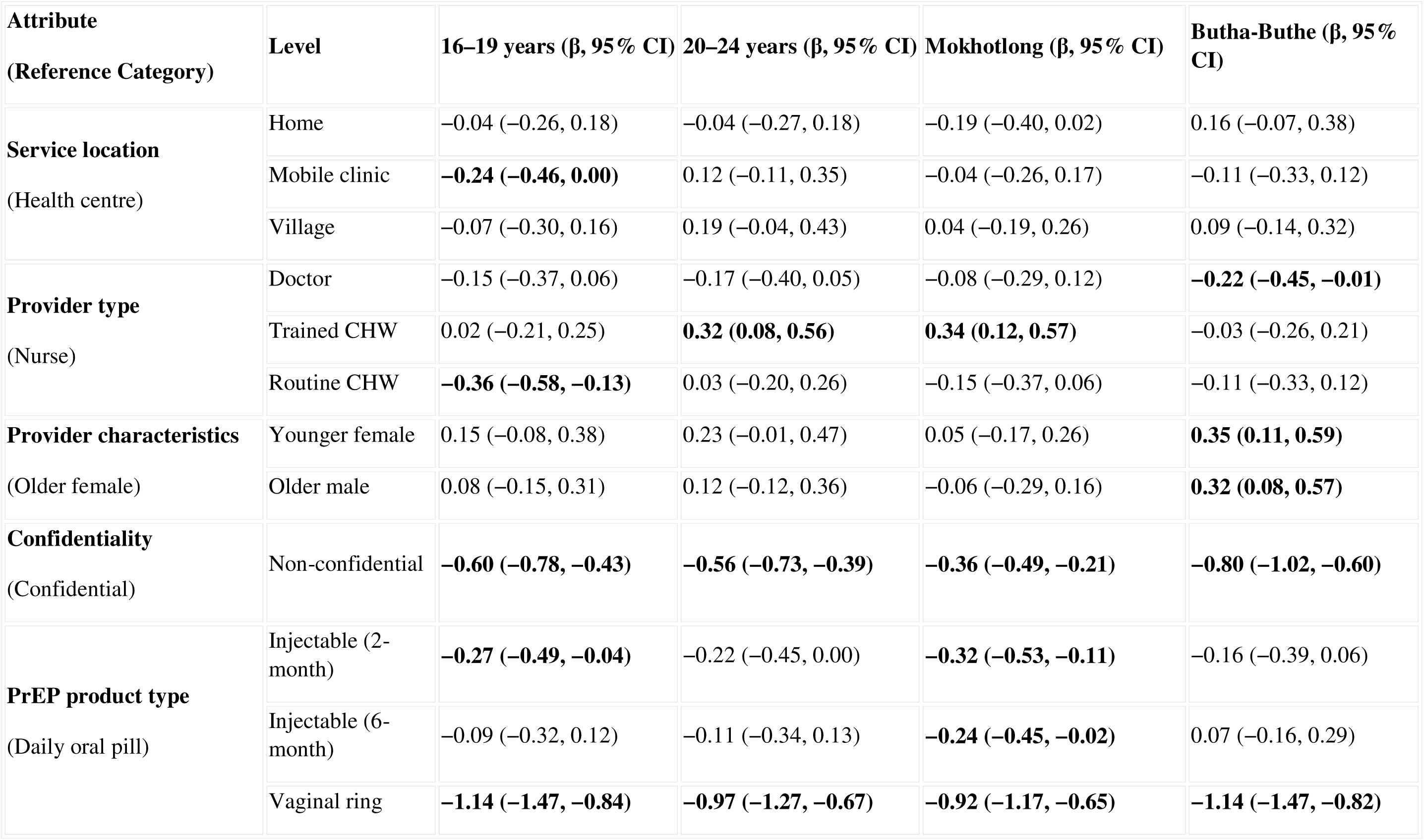
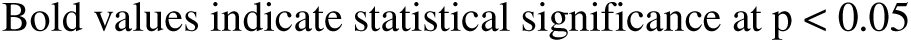
Subgroup analysis of PrEP service delivery preferences by age group and district.

**Table 3b.**
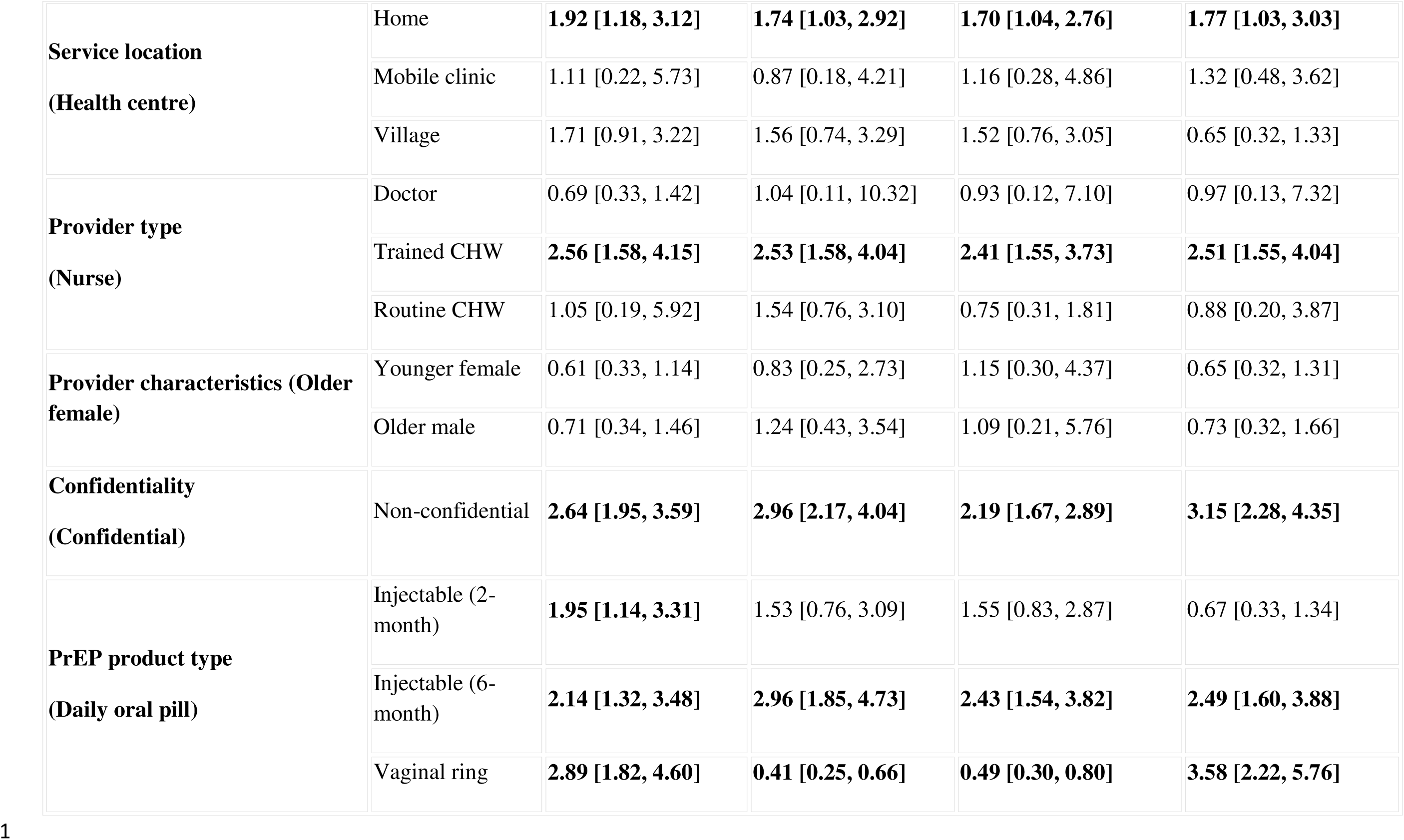
Subgroup analysis of PrEP service delivery preferences by age group and district: random coefficients.

Models with two to four classes were compared, and a two-class model was selected based on model fit and classification accuracy (mean posterior probability 0.84; Supplementary Table S5). The final model (n = 537) identified two distinct preference profiles: **Class 1** (n = 282; 53%) and **Class 2** (n = 255; 47%) (Figure 3).

**Figure 3:**
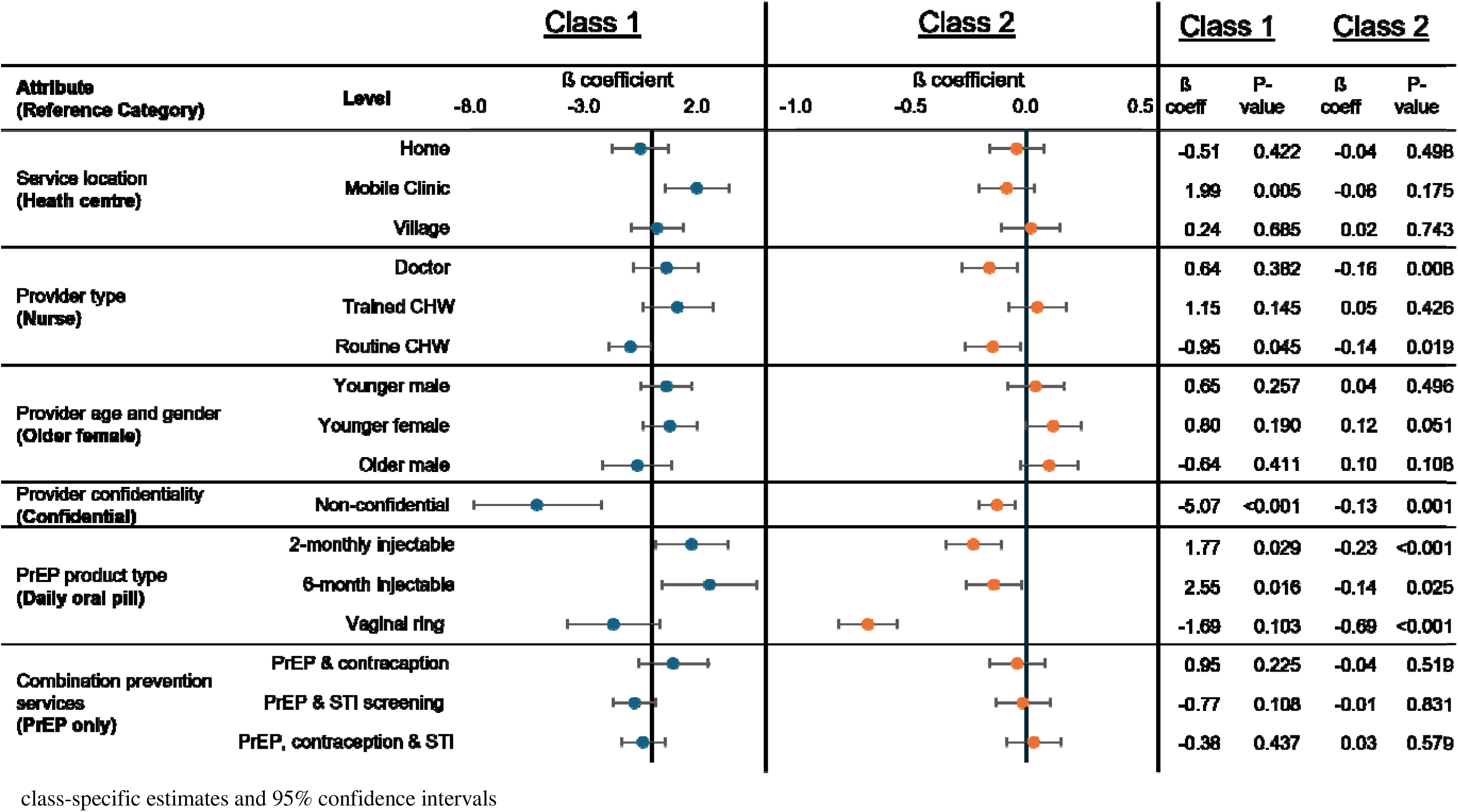
Latent class analysis of PrEP service delivery preferences.

**Class 1 (53%)** showed stronger and more clearly differentiated preferences across attributes. Mobile clinic delivery was preferred (β = 1.99; p = 0.005), while services delivered by routine CHWs were less preferred (β = −0.95; p = 0.045). Provider characteristics were not associated with choice

Provider confidentiality had a strong influence, with substantial aversion to non-confidential providers (β = −5.07; p < 0.001). PrEP product type also influenced preferences, with both the 2-monthly injectable (β = 1.77; p = 0.029) and 6-monthly injectable PrEP (β = 2.55; p = 0.016) preferred over the daily oral pill, while no preference was observed for the vaginal ring. Additional preventive services were not associated with choice.

**Class 2 (47%)** showed a more moderate pattern of preferences. No preferences were observed for service location. Services provided by doctors (β = −0.16; p = 0.008) and routine CHWs (β = −0.14; p = 0.019) were less preferred compared with nurses, while other provider types were not associated with choice. Provider characteristics were not associated with choice.

Provider confidentiality remained significant, with non-confidential providers less preferred (β = −0.13; p = 0.001). PrEP product type also influenced preferences, with the vaginal ring (β = −0.69; p < 0.001), 2-monthly injectable (β = −0.23; p < 0.001), and 6-monthly injectable PrEP (β = −0.14; p = 0.025) all less preferred compared with the daily oral pill. Additional preventive services were not associated with choice.

## Discussion

This is the first DCE among AGYW living in remote villages in Lesotho to assess preferences for PrEP service delivery and trade-offs across multiple attributes within available service options. The main effects model used the standard of care as the reference to compare preferences with current service delivery. Across all models, provider confidentiality emerged as the strongest and most consistent driver of choice and was not easily traded off against other service features. Preferences for other attributes were only evident when services were described as confidential, suggesting that they became meaningful only once confidentiality was assured. Confidentiality concerns are a well-established barrier to PrEP uptake and continuation among AGYW [10,26] and should be considered a minimum standard in the design of PrEP services. Without confidentiality, expanding service delivery options alone is unlikely to increase uptake.

Overall, facility-based care and daily oral PrEP were preferred; however, participants showed clear preferences for certain provider types and characteristics, particularly trained CHWs and younger female providers, while services provided by doctors were less preferred. These findings support task shifting, suggesting that CHWs can expand access and reduce the burden on nurses without reducing demand for services. While the current standard of care is broadly aligned with these preferences, targeted adjustments may further improve acceptability and access. These include expanding delivery through trained and supported CHWs and aligning provider characteristics with AGYW preferences, particularly by allowing for female providers where possible.

Preferences for PrEP product type were more varied. Daily oral PrEP was generally preferred, the vaginal ring was least preferred, and preferences for long-acting injectables were mixed. Within the context of current service delivery, daily oral PrEP remained the most acceptable option overall. However, interest in long-acting injectable PrEP, particularly in one preference group identified in the latent class analysis, suggests openness to additional options as they become available and that injectable formulations may increase demand for some AGYW. Previous studies emphasise the importance of choice in HIV prevention [4,16]. Our findings support this, but also indicate that not all options are equally acceptable. In this setting, and given resource constraints, it may be more appropriate to prioritise daily oral PrEP alongside long-acting injectables. In contrast, the consistently low acceptability of the vaginal ring suggests that it is unlikely to be widely used in this setting.

Service delivery preferences differed across age groups and districts in both stratified and latent class analyses. Younger AGYW were less open to mobile clinic services and routine CHW delivery, while older AGYW showed a preference for trained CHWs. Younger adolescents often face greater barriers to sustained engagement in HIV prevention, including stigma, lower agency, and logistical constraints [28], which may contribute to these differences and suggest that community-based delivery may be more acceptable for older AGYW. These findings highlight the importance of age when designing service delivery models, rather than assuming a single approach will meet the needs of all AGYW.

District-level differences further support a context-specific approach. In Butha-Buthe, preferences were more strongly influenced by provider characteristics, whereas in Mokhotlong, trained CHWs were preferred alongside lower acceptance of injectable PrEP formulations. These patterns may reflect variation in geographic access and service delivery environments, with more remote communities in Mokhotlong likely relying more on community-based care and established relationships with CHWs. Together, these findings support the decentralisation of PrEP services as a strategy to improve access, particularly in remote settings. Mobile and community-based delivery models, including CHWs, may offer a practical way to reach AGYW who face barriers to facility-based care. However, variation in preferences suggests that these approaches should be implemented flexibly, considering differences by age, context, and product type.

Combination prevention services, including contraception and STI screening, were not significant drivers of choice, suggesting that they were less influential than other attributes. This may reflect that these features were secondary to more immediate concerns, such as confidentiality and provider characteristics, and were therefore more easily traded off. Accessing multiple services in a single visit may also introduce barriers, including longer waiting times [29] or increased visibility, which could heighten concerns around stigma. Despite this, combination prevention remains an important component of comprehensive HIV prevention and should continue to be offered as part of PrEP service delivery.

Variation in preferences across individuals and subgroups suggests that a single delivery model is unlikely to meet the needs of all AGYW. In practice, this may involve offering more than one way to access PrEP, rather than relying on a single approach. Findings from the latent class analysis suggest that some AGYW may be open to community-based delivery, including mobile clinics, alongside interest in long-acting injectable PrEP, while others show more moderate preferences that align with the current standard of care. This supports offering different delivery approaches in parallel to better reflect how AGYW engage with services in this setting.

This study has several strengths. Data were collected during routine community health activities, supporting the practical relevance of the findings. The inclusion of AGYW living in remote areas of Lesotho, alongside a high response rate and population-based design, strengthens the representativeness of the results. Attributes were informed by qualitative research and local stakeholder input, and the inclusion of emerging PrEP products, such as lenacapavir, allowed the study to capture forward-looking preferences.

Several limitations should be noted. As a stated-preference method, the DCE reflects hypothetical choices and may not translate directly into PrEP uptake. The forced-choice design did not include an opt-out option, which may overstate preferences. The provider confidentiality attribute included clearly directional levels, reflecting strong prioritisation rather than direct comparability with other attributes. Data were self-reported and collected by CHWs, which may introduce social desirability or interviewer bias

Finally, as the study was conducted in two remote districts of Lesotho, the findings may not be generalisable to other settings, including urban or peri-urban areas.

### Conclusions

The current standard of care for PrEP delivery in Lesotho is broadly consistent with the preferences of AGYW, but there are clear opportunities to improve access and acceptability. Decentralising PrEP delivery, particularly through trained and supported CHWs and community-based models, may help reach AGYW living in remote settings and was generally well received. Across all models, provider confidentiality emerged as a non-negotiable requirement and should be prioritised in programme design.

Preferences for PrEP products were more varied. Daily oral PrEP remained the most acceptable option overall, with some interest in long-acting injectables, while the vaginal ring was consistently least preferred. Rather than expanding all available options, efforts may be better focused on delivering PrEP in ways that are accessible, confidential, and acceptable to AGYW.

## Data Availability

All data produced in the present study are available upon reasonable request to the authors

## Authors’ contributions

AW and MS conceptualised the study. AW, MS, TC, and GG developed the methodology. AW, OP, and MS conducted the formal analysis. GSS and ES were responsible for data management. RG, MM, LM, MCM, MPS, MK, PG, and IA supported data collection and study implementation. NDL, AA, MW, TC, and GG provided supervision. AW drafted the original manuscript. All authors contributed to manuscript review and editing and approved the final version.

## Acknowledgements

The authors thank the study participants and community health workers for their time and contributions. We also acknowledge the Lesotho Ministry of Health and the ComBaCaL team for their support in study implementation and data collection.

## Data Availability Statement

The datasets used and analysed during the current study are available from the corresponding author upon request.

## Supporting Information

Supplementary materials, including example DCE choice tasks and additional study materials, are available from the corresponding author upon request:

- Table S1a–S1b. Definitions and levels of DCE attributes included in the experimental design
- Table S2. Validity and reliability checks of DCE responses
- File S1. Full DCE experimental design matrix
- File S2. Dominant and inferior (repeated) choice set used for internal consistency
- File S3. Example of a DCE choice set presented to participants
- File S4. Binary-coded dataset used for model estimation

## List of abbreviations

AIC: Akaike information criterion
AGYW: Adolescent girls and young women
BIC: Bayesian information criterion
CHT□: Community Health Toolkit
CHW: Community Health Worker
ComBaCaL: Community-Based Chronic Care Lesotho
DCE: Discrete Choice Experiment
HIV: Human immunodeficiency Virus
PrEP: Pre-Exposure Prophylaxis
STI: Sexually transmitted infections

