## Supplementary Tables S1a and S1b for "Preferences for PrEP service delivery among adolescent girls and young women in remote villages in Lesotho: a discrete choice experiment"

**Supplementary Table S1a:** Overview and definitions of DCE attributes included in the design of hypothetical PrEP service delivery options.

| **Service Location** | A location/place where you can receive healthcare services or collect medication |
| --- | --- |
| **Provider Type** | A person who is educated or trained to provide healthcare services |
| **Provider Characteristics** | Age in years and sex (male or female) of a healthcare provider |
| **Provider Confidentiality** | A person’s way of thinking, feeling, or behaving and whether they can be trusted with personal information |
| **PrEP product** | A type of product with a particular shape, size and mode of administration |
| **Combiation of additional prevention Services** | Products or services that can be offered alone or together |

**Supplementary Table S1b.** List of levels and corresponding descriptions for each DCE attribute used in the final experimental design.

|  | **Level** | **Label** | **Definition/description** |
| --- | --- | --- | --- |
| **Service Location** | 1 | Home (in the village) | At the home in the village where you live |
|  | 2 | Healthcare facility | At the nearest clinic or hospital to where you live |
|  | 3 | Mobile clinic | At the trailer/van that provides healthcare services near the village where you live |
|  | 4 | In the village (but not at home) | At a location near to your village (e.g., near the well/tap, by a soccer field), but not at your home |
| **Provider** **Type** | 1 | Doctor | A person with a medical degree qualified by graduation from an accredited medical school with a national license to practice as a doctor |
|  | 2 | Nurse | A nursing professional qualified by graduation from an accredited school of nursing with a national license to practice as a nurse |
|  | 3 | CHW (ComBaCaL) | A community-elected lay health-worker assisted by an eHealth application to provide healthcare in villages, trained and supervised by an NGO |
|  | 4 | CHW (Routine) | A community-elected lay health-worker employed by the Ministry of Health who provide healthcare services in villages, trained and supervised by the Ministry of Health |
| **Provider Characteristics** | 1 | Younger male  (<30 years) | A male younger than 30 years |
|  | 2 | Younger female  (<30 years) | A female younger than 30 years |
|  | 3 | Older male  (>30 years) | A male 30 years or older |
|  | 4 | Older female  (>30 years) | A female 30 years or older |
| **Provider**  **Confidentiality** | 1 | Non-confidential | Is known to have conversations or report about other people that involves information that may or may not be true |
|  | 2 | Confidential | Does not have conversations or reports about other people that involves information that may or may not be true |
| **PrEP Product** | 1 | Oral pill (taken daily) | A blue, oval-shaped pill containing anti-retroviral medication that is taken every day, by mouth, to prevent HIV infection |
|  | 2 | Injection in buttocks (taken every 2 months) | A syringe and needle containing a clear liquid with antiretroviral medication that is injected into the buttocks every 28 days to prevent HIV infection |
|  | 3 | Injection in abdomen/stomach (every 6 months) | A syringe and needle containing a clear liquid with antiretroviral medication that is injected into the abdomen every 6 months to prevent HIV infection |
|  | 4 | Vaginal ring (vaginal insertion, replaced every month) | A silicone ring containing anti-retroviral medication that is inserted into the vagina for 28 days to prevent HIV infection |
| **Combination of additional prevention services** | 1 | PrEP only | A product that contains anti-retroviral medication only |
|  | 2 | PrEP & Contraception | A Product/s that contain anti-retroviral medication and contraception only |
|  | 3 | PrEP & STI screening | A Product/s that has anti-retroviral medication and includes STI screening only |
|  | 4 | PrEP, Contraception & STI screening | A Product/s that contains anti-retroviral medication, contraception and includes STI screening |

|  | **Level** | **Label** | **Definition/description** |
| --- | --- | --- | --- |
| **Service Location**  Definition: A location/place where you can receive healthcare services or collect medication | 1 | Home (in the village) | At the home in the village where you live |
|  | 2 | Healthcare facility | At the nearest clinic or hospital to where you live |
|  | 3 | Mobile clinic | At the trailer/van that provides healthcare services near the village where you live |
|  | 4 | In the village (but not at home) | At a location near to your village (e.g., near the well/tap, by a soccer field), but not at your home |
| **Provider** **Type**  Definition: A person who is educated or trained to provide healthcare services | 1 | Doctor | A person with a medical degree qualified by graduation from an accredited medical school with a national license to practice as a doctor |
|  | 2 | Nurse | A nursing professional qualified by graduation from an accredited school of nursing with a national license to practice as a nurse |
|  | 3 | CHW (ComBaCaL) | A community-elected lay health-worker assisted by an eHealth application to provide healthcare in villages, trained and supervised by an NGO |
|  | 4 | CHW (Routine) | A community-elected lay health-worker employed by the Ministry of Health who provide healthcare services in villages, trained and supervised by the Ministry of Health |
| **Provider Characteristics**  Definition: Age in years and sex (male or female) of a healthcare provider | 1 | Younger male  (<30 years) | A male younger than 30 years |
|  | 2 | Younger female  (<30 years) | A female younger than 30 years |
|  | 3 | Older male  (>30 years) | A male 30 years or older |
|  | 4 | Older female  (>30 years) | A female 30 years or older |
| **Provider** **Confidentiality**  Definition: A person’s way of thinking, feeling, or behaving and whether they can be trusted with personal information | 1 | Non-confidential | Is known to have conversations or report about other people that involves information that may or may not be true |
|  | 2 | Confidential | Does not have conversations or reports about other people that involves information that may or may not be true |
| **PrEP Product**  Definition: A type of product with a particular shape, size and mode of administration | 1 | Oral pill (taken daily) | A blue, oval-shaped pill containing anti-retroviral medication that is taken every day, by mouth, to prevent HIV infection |
|  | 2 | Injection in buttocks (taken every 2 months) | A syringe and needle containing a clear liquid with antiretroviral medication that is injected into the buttocks every 28 days to prevent HIV infection |
|  | 3 | Injection in abdomen/stomach (every 6 months) | A syringe and needle containing a clear liquid with antiretroviral medication that is injected into the abdomen every 6 months to prevent HIV infection |
|  | 4 | Vaginal ring (vaginal insertion, replaced every month) | A silicone ring containing anti-retroviral medication that is inserted into the vagina for 28 days to prevent HIV infection |
| **Combination of additional prevention services**  Definition: Products or services that can be offered alone or together | 1 | PrEP only | A product that contains anti-retroviral medication only |
|  | 2 | PrEP & Contraception | A Product/s that contain anti-retroviral medication and contraception only |
|  | 3 | PrEP & STI screening | A Product/s that has anti-retroviral medication and includes STI screening only |
|  | 4 | PrEP, Contraception & STI screening | A Product/s that contains anti-retroviral medication, contraception and includes STI screening |
