## Supplementary Table S5 for "Preferences for PrEP service delivery among adolescent girls and young women in remote villages in Lesotho: a discrete choice experiment"

**Supplementary Table S5.** Comparison of model fit statistics for two-, three-, and four-class latent class models

| **Number of classes** | **Log likelihood** | **AIC** | **BIC** |  |  |  | **Mean probability of class membership** |
| --- | --- | --- | --- | --- | --- | --- | --- |
| **2 classes** | -2712.6 | 5491.2 | 5701.3 |  |  |  | 0.84 |
| **3 classes** | -2689.7 | 5480.0 | 5797.7 |  |  |  | N/A* |
| **4 classes** | -2654.5 | 5443.0 | 5869.5 |  |  |  | N/A* |

*For the three- and four-class models, class separation was poor (collapsed or non-distinct classes).
